# The HIV-treatment-as-prevention adoption cascade among U.S. men and gender-minority individuals who have sex with men

**DOI:** 10.64898/2026.01.19.26344396

**Authors:** Étienne Meunier, Daniel Sauermilch

## Abstract

HIV treatment can suppress viral load and prevent transmission between sex partners, a strategy known as treatment as prevention (TasP). TasP is key for ending the HIV epidemic, and it is important to understand its adoption among priority populations. We examined the TasP adoption cascade using cross-sectional survey data from 1443 U.S. men and transgender, gender-nonconforming, and nonbinary individuals who reported having sex with men. Most participants (82.4%; n = 1189/1443) reported prior awareness of TasP, but only 52.6% of them (n = 625/1189) perceived it as effective at preventing HIV transmission. Of those, 83.8% (n = 524/625) indicated being willing to rely on TasP, among whom 30.2% (n = 158/524) reported having recently done so. Among participants aware of TasP, we compared those who perceived it as effective to those who did not. Participants who did not have HIV and never used PrEP were less likely to agree with TasP’s effectiveness than those who had used PrEP or had HIV. Those who had learned about TasP from a sex partner or who had a partner of different HIV status were more likely to perceive it as effective. TasP promotion appears to have achieved broad awareness, but future efforts should aim at increasing the understanding of its effectiveness, especially among those not connected to HIV-related services, organizations, or communities.

**Public Health Significance:** TasP is an important tool to end the HIV epidemic. Examining stages of its adoption can inform tailored promotion among priority populations. In our study, many participants were aware of TasP, but fewer perceived it as effective. Monitoring uptake over time will allow for responsive promotion strategies as attitudes continue to evolve.

## Introduction

Clinical trials show that people with HIV who achieve and maintain viral suppression do not transmit the virus to their sex partners,^1,2^ a strategy known as “treatment as prevention” (TasP) and promoted under the slogan “Undetectable equals untransmittable” (U=U). Promoting TasP can reduce HIV-related stigma, remove barriers to HIV testing, and encourage engagement in HIV care and treatment adherence.^3,4^

TasP promotion is particularly important in populations with high HIV prevalence and incidence, including gay, bisexual, and other men who have sex with men (MSM) and transgender, gender nonconforming, and nonbinary (TGNCNB) individuals. In 2022, more than 800,000 men in the United States were living with HIV, and about two thirds of new diagnoses were attributed to male-to-male sexual contact; TGNCNB individuals were also disproportionately affected by HIV.^5^

Since the launch of the U=U campaign in 2016, surveys that included U.S. sexual– or gender-minority people have found that most respondents were aware of TasP but fewer agreed it would effectively prevent HIV transmission, and even fewer were willing to rely on it.^4,6–8^ In a 2017–2018 survey with 3268 HIV-negative MSM and TGNCNB individuals, 85% were aware of TasP, but only 42% of those trusted its effectiveness, and far fewer reported willingness to have condomless sex with a partner with suppressed HIV.^8^

Although some studies have examined individual components of TasP acceptability, few assessed the full continuum of adoption. Drawing from the Precaution Adoption Process Model^9^ the stages of TasP adoption can be conceptualized as a cascade: gaining awareness of TasP, agreeing with its effectiveness, being willing to rely on it, and using it. The cascade framework has been used in HIV research to look at adoption of PrEP or HIV treatment.^10,11^ The goal of this study was to map the TasP adoption cascade in a survey of 1443 U.S. MSM and TGNCNB individuals who have sex with men. Identifying which stages represent the main breakpoints in the cascade can inform the focus of future TasP promotion efforts.

## Methods

### Participants

We conducted a cross-sectional online survey between December 2020 and June 2021. Participants could self-enroll through advertisements on social media (Facebook, Instagram, and Reddit) or dating/hookup applications (Grindr, GROWLr, and Scruff). Ads indicated that researchers at Columbia University were conducting a study on the sexual health of men and TGNCNB individuals who have sex with men, and that eligible participants who completed the survey would be able to enter a raffle for a $200 gift card.

The ads directed interested participants to the survey page on Qualtrics where, after informed consent, they were asked a few questions to determine eligibility. To proceed to the full survey, respondents had to report: a) being at least 18 years old; b) living in the United States; c) fluency in English; d) identifying as a cisgender man or as transgender, gender nonconforming, or nonbinary; and e) having had sex with a male partner in the previous 12 months. The online survey was initiated 1996 times and 1443 unique participants were eligible and completed the survey.

### Measures

The survey elicited demographics and information about sexual health and behaviors (detailed in Table 1). We assessed TasP acceptability using a series of questions (included as supplemental material) we developed based on our review of the relevant literature and prior work on TasP.^12,13^ For the current report, we looked at four stages of TasP adoption measured as follows:

1) *awareness*: reported having heard of TasP prior to the survey;
2) *perceived effectiveness*: said there would be very low or no risk of HIV transmission if a man with undetectable HIV had anal sex as the insertive (top) partner with an HIV-negative man without using condoms or PrEP;
3) *willingness to use*: reported willingness to have condomless sex with a partner of a different HIV status if the individual with HIV had an undetectable viral load;
4) *recent use*: HIV-negative participants who reported condomless sex with partners with undetectable HIV in the prior 6 months, or participants with undetectable HIV who did so with HIV-negative partners who were not using PrEP.

**Table 1.**
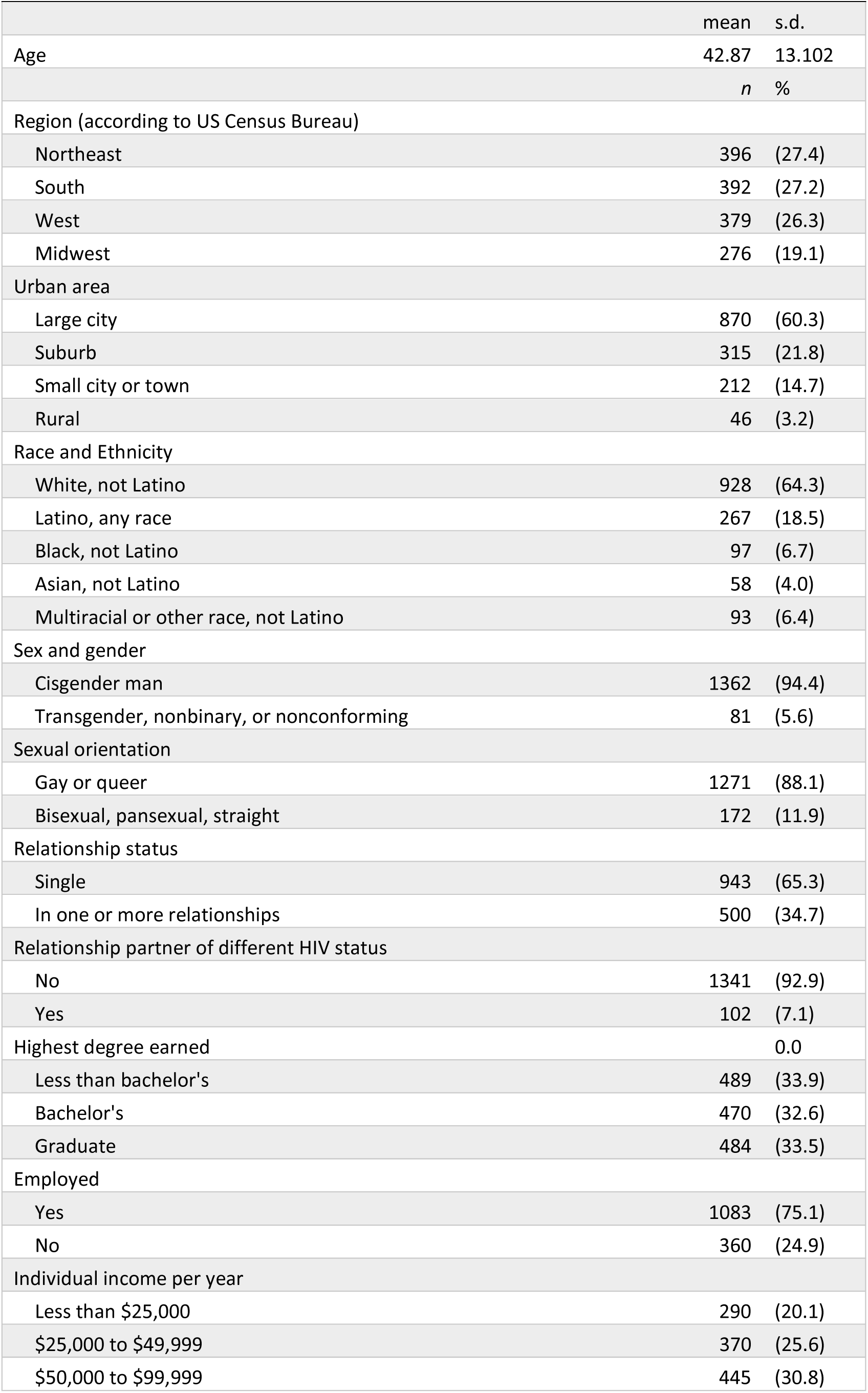
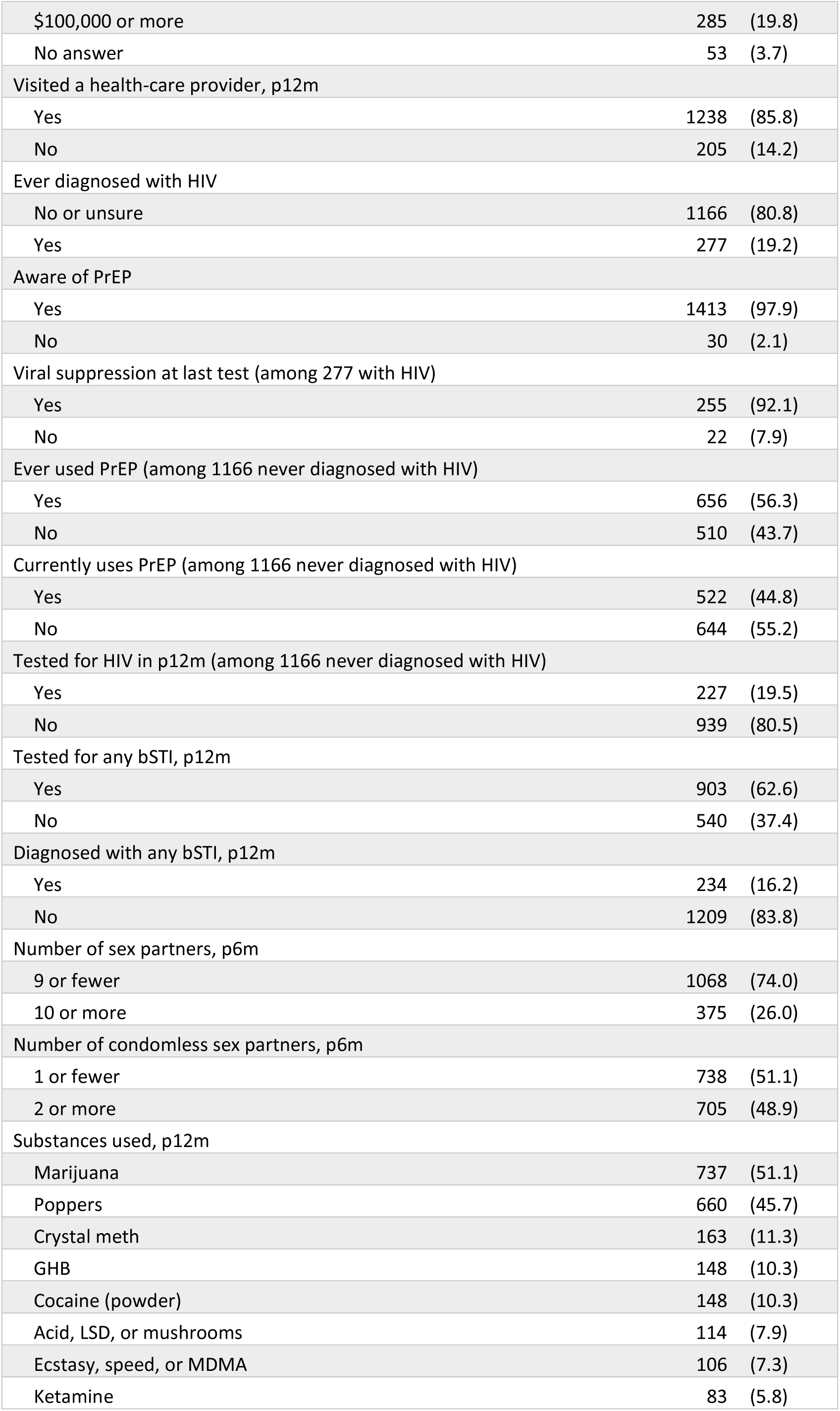
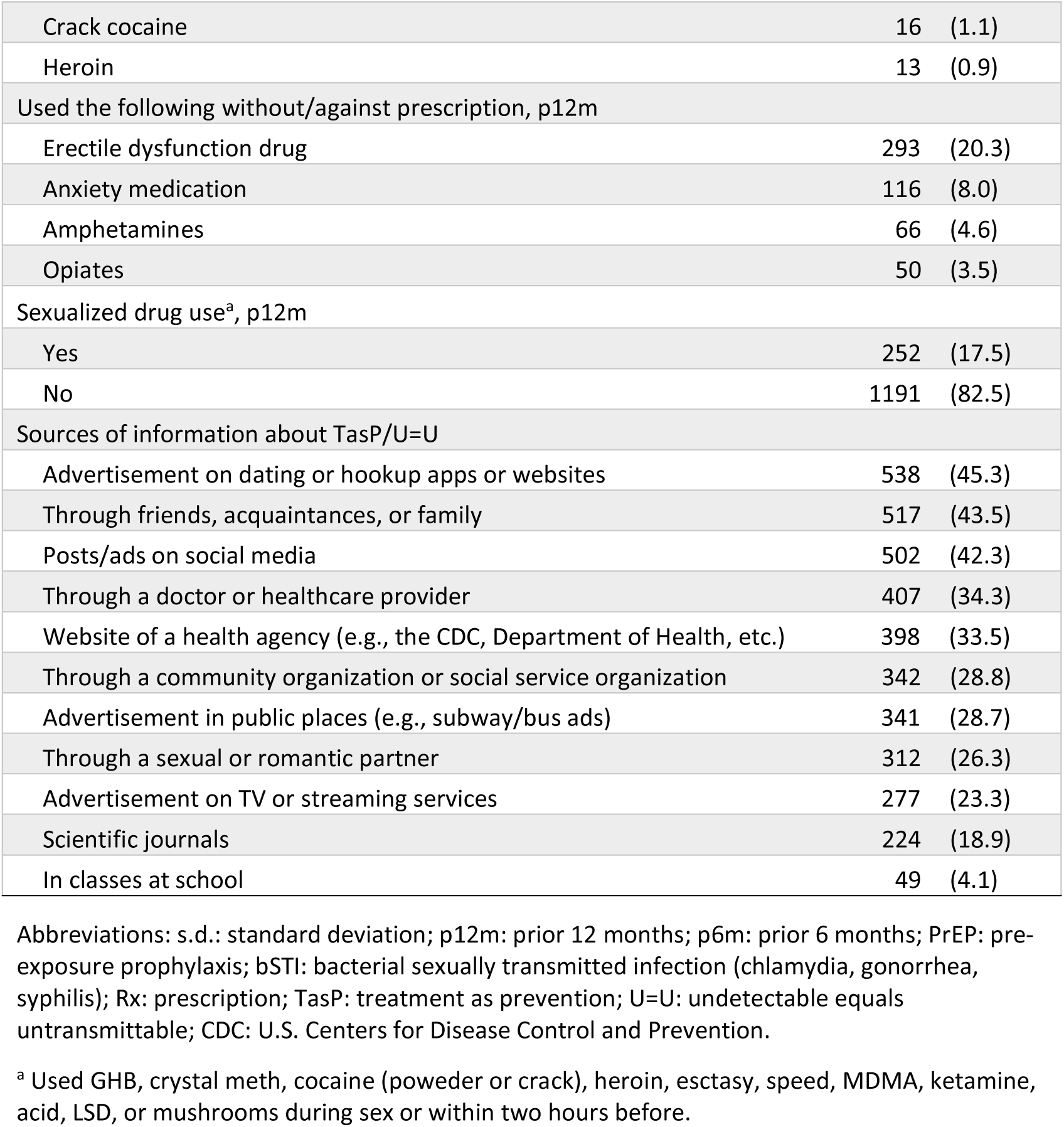
Participant characteristics and descriptive statistics.

### Analysis

We used descriptive statistics to characterize participants and categorize them along the stages of the TasP adoption cascade. We identified the main breakpoint across the first three stages by examining retention percentages.^14^ We compared participants across the two stages around the main breakpoint using bivariate binary logistic regressions to assess associations between demographic, behavior, and health variables (α = .1). Variables significant at this level were then included in a multivariable binary logistic regression (α = .05). Analyses were conducted using SPSS version 28.

## Results

Participants’ characteristics are displayed in Table 1. The mean age was 42.9 years old. Participants represented various U.S. regions, though most (60.3%; n = 870) said they resided in large cities and only 3.2% in rural areas. Most (64.3%; n = 928) identified as White and not Latino, gay or queer (88.1%; n = 1271), and cisgender men (94.4%; n = 1362). About two thirds (66.1%; n = 954) had earned a bachelor’s degree and 65.3% were single. About one fifth of participants (19.2%; n = 277) reported having been diagnosed with HIV and, of them, 92.1% (n = 255) said they had reached viral suppression at their last viral load test. Among participants who reported having never been diagnosed with HIV, 56.3% (n = 656) had ever used PrEP and 44.8% (n = 522) were currently using it.

Figure 1 illustrates the cascade of TasP adoption across four stages. Awareness (Stage 1) was high, with 82.4% (1189/1443) reporting prior knowledge of TasP. However, nearly half did not progress to Stage 2, as only 52.6% (625/1189) of those aware perceived TasP as effective. Attrition was minimal to Stage 3, as 83.8% (524/625) of those who perceived effectiveness were also willing to rely on TasP. At Stage 4, 30.2% (158/524) reported recent reliance on TasP, an expected decline given the six-month reference period, the exclusion of TasP use with condoms, and the fact that only participants with a known serodifferent partner could report its use. Accordingly, the most meaningful breakpoint in the cascade was between awareness (Stage 1) and perceived effectiveness (Stage 2).

**Figure 1.**
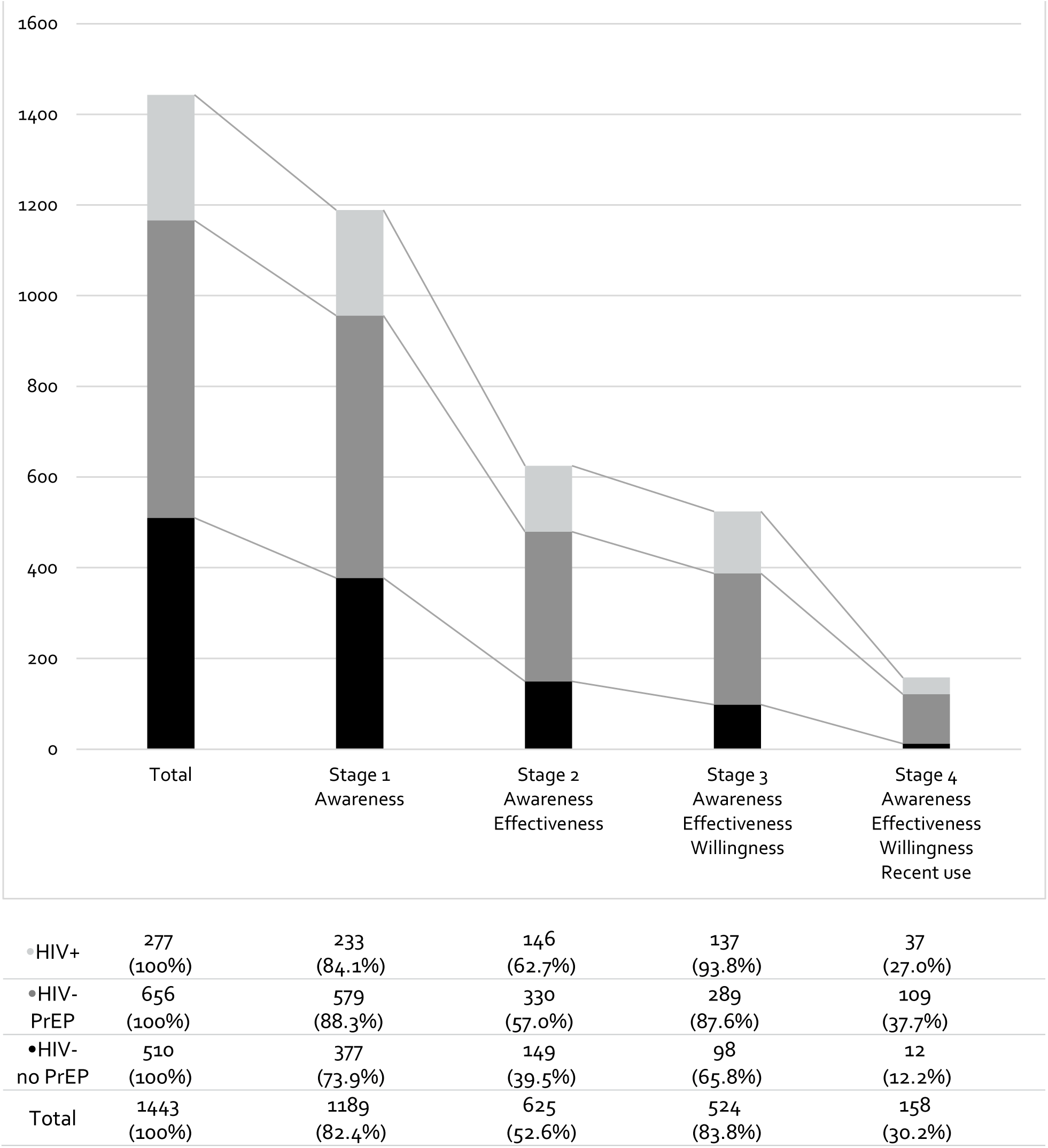
TasP acceptability cascade. The main drop in the TasP adoption cascade was between becoming aware of the strategy and perceiving it as effective. Percentages are among participants who reached the prior stage. Recent use refers to the prior six months and excludes participants who reported unsuppressed HIV.

Across every stage, declines were most pronounced among participants who reported never having been diagnosed with HIV and never having used PrEP. For example, among those aware of TasP, only 39.5% of this group perceived it as effective, compared with 57.0% of participants who reported no HIV diagnosis but had used PrEP, and 62.7% of those who reported an HIV diagnosis.

We compared participants who were aware of TasP but did not perceive it as effective (n = 564) to those who were both aware and perceived it as effective (n = 625) (Table 2). Increased age was associated with lower agreement with TasP’s effectiveness (aOR = 0.98; 95% CI: 0.97–0.99). Compared to White non-Latino participants, those identifying as Latino of any race (aOR = 0.69; 95% CI: 0.49–0.96) or as multiracial or of other race (aOR = 0.40; 95% CI: 0.24–0.67) were less likely to perceive TasP as effective. Perceived effectiveness was higher among participants who were in serodifferent relationships (aOR = 1.74; 95% CI: 1.05–2.86) and those who had completed a bachelor’s or higher degree (aOR = 1.46; 95% CI: 1.11–1.92).

**Table 2.**
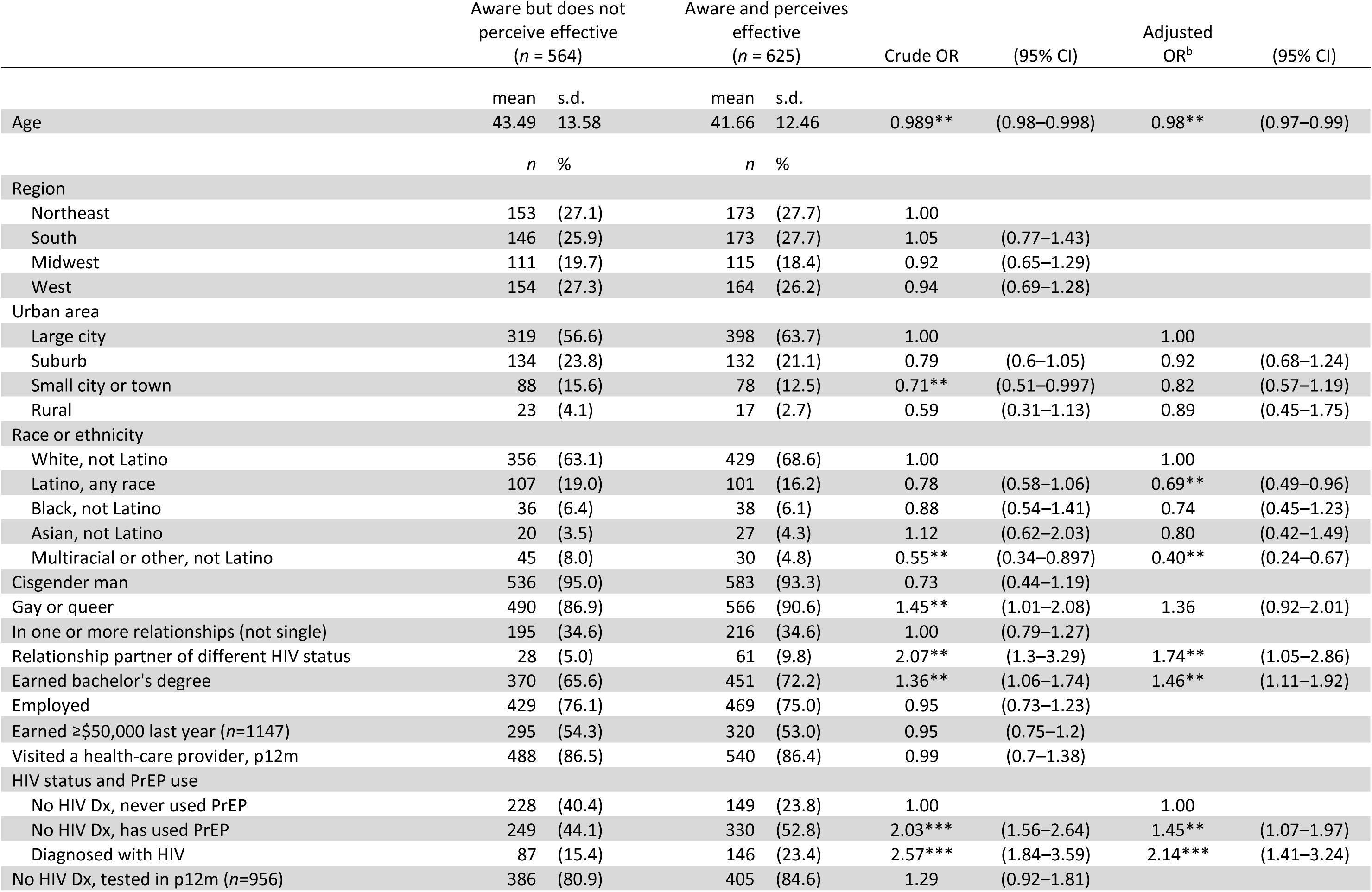
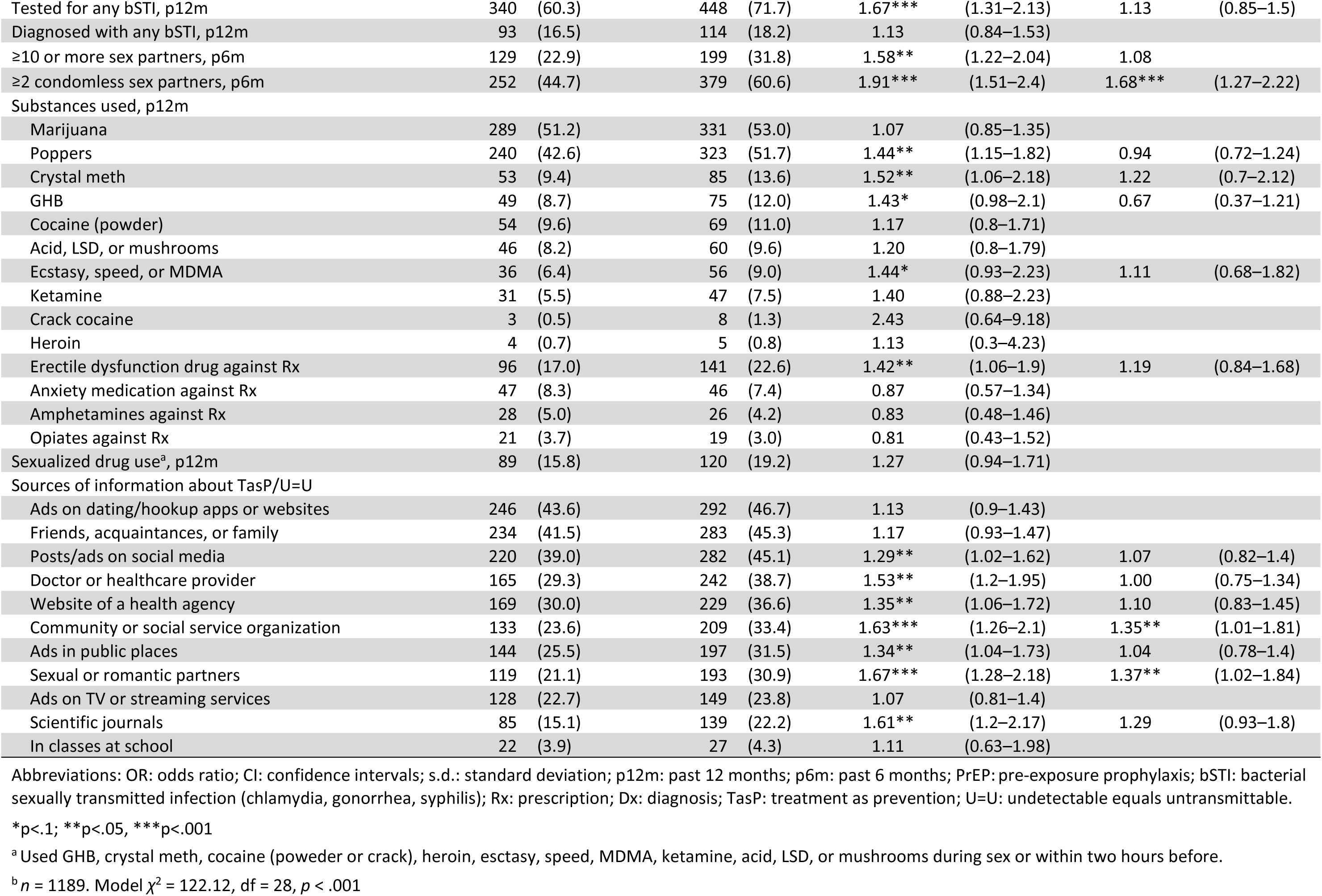
Logistic regression models comparing participants at stages 1 and 2 of TasP adoption.

Compared to participants who reported no HIV diagnosis and never using PrEP, those who had used PrEP (aOR = 1.45; 95% CI: 1.07–1.97) and those who had HIV (aOR = 2.14; 95% CI: 1.41–3.24) were more likely to perceive TasP as effective. Participants reporting two or more condomless sex partners in the prior six months were also more likely to perceive TasP as effective (aOR = 1.68; 95% CI: 1.27–2.22). Finally, participants who reported learning or receiving information about TasP/U=U from community organizations (aOR = 1.35; 95% CI: 1.01–1.81) or from sex or romantic partners (aOR = 1.37; 95% CI: 1.02–1.84) were more likely to perceive TasP as effective.

## Discussion

Our survey mapped the stages of TasP adoption among 1443 U.S. men and TGNCNB individuals who have sex with men. While most participants (82.4%) reported prior awareness of TasP, only about half of them (52.6%) also agreed that it was effective at preventing HIV transmission, marking the main breakpoint in the adoption cascade. These findings suggest that, when these data were collected, awareness-raising efforts had been successful, but understanding of TasP’s effectiveness was lagging.

The high awareness and lower perceived effectiveness we observed align with findings from other surveys among MSM or TGNCNB individuals in the United States.^4,7,8^ This pattern may reflect how early TasP promotion efforts prioritized broad messaging (such as social marketing campaigns featuring the U=U slogan) over detailed education about the strategy. For instance, an analysis of U.S. health departments’ websites found that many only offered vague, incomplete, or inaccurate information about TasP.^15^ Another study among men with HIV and healthcare providers in the United States and Australia found that providers were often hesitant to educate patients about TasP, or did so ambiguously or inaccurately.^16^ While TasP promotion appears to have succeeded in raising awareness, additional efforts may be needed to ensure people fully understand and trust its effectiveness. To inform such efforts, we compared participants across these two stages of TasP adoption.

Compared to participants who reported no HIV diagnosis and never having used PrEP, those who had HIV or had used PrEP were significantly more likely to agree that TasP was effective, a pattern consistent with other surveys.^4,6–8^ Among HIV-negative individuals, those with PrEP experience may be more engaged with sexual-health services, where they can access more detailed information about TasP. Supporting this interpretation, participants who reported learning about TasP from a community organization were also more likely to perceive it as effective. Barriers to PrEP and TasP adoption may also overlap; for example, medical mistrust is a known barrier to PrEP,^17^ and skepticism about the ability of medications to prevent HIV may similarly shape perceptions of TasP.

For people with HIV, TasP is a key strategy for preventing transmission to partners, and they may be more likely to seek information about it or receive education from providers. However, even though participants with HIV were the most likely of all three groups we examined to perceive TasP as effective, nearly half of them did not. Prior qualitative research with individuals in serodifferent relationships highlighted worries that, despite viral suppression, some residual risk of transmission might remain.^18,19^ These studies predated the launch of the U=U campaign, which aimed to reduce HIV stigma and improve uptake of treatment among people with HIV. Examining the reasons why a substantial proportion of people with HIV today remain unconvinced of TasP’s effectiveness will be important to guide future education efforts.

Participants who reported learning or receiving information about TasP from a sex partner were more likely to agree with its effectiveness, along with those who reported having a relationship partner of a different HIV status. Prior studies had discussed how TasP could facilitate relationships across serostatus^18^ and bridge the “serodivide” between people with and without HIV.^20^ While our data cannot determine whether TasP understanding leads to serodifferent partnerships or vice versa, it is likely that such experiences prompt deeper engagement with the strategy. For example, participants in a prior qualitative study described increasingly trusting TasP as they kept using it.^12^ For individuals without serodifferent partners, TasP may remain an abstract concept they feel no need to understand in depth. Interventions that foster dialogue across HIV status, whether through peer education, provider support, or community programming, may help increase TasP adoption.

We found that participants who identified as Latino or as multiracial were less likely to perceive TasP as effective than those who identified as White, with no significant differences for Black-identifying participants; a pattern similar to that observed in a prior U.S. survey.^8^ The absence of statistical significance might be attributable to the difficulty of enrolling sufficiently large numbers of Black MSM and TGNCNB individuals to participate in surveys, and should not be interpreted as evidence of no difference. Studies seem to consistently show that there are racial/ethnic disparities in adoption of biomedical HIV prevention, which should be addressed considering how Black and Latino individuals in the United States are disproportionately affected by HIV.^5^ The low perceived effectiveness of TasP among MSM and TGNCNB individuals of color may reflect medical mistrust; however, such skepticism should not be viewed as a lack of information, but rather as a response to a broader history of exclusionary clinical practices, discrimination in medical settings, and inequitable access to care.^21^

Most participants (83.8%) who agreed with TasP’s effectiveness also reported willingness to rely on the strategy, representing the third stage of adoption. In a prior qualitative study,^12^ some participants expressed being unwilling to use TasP despite perceiving it as effective, citing deep-rooted fears about HIV or the difficulty to know a partner’s viral load. While such barriers warrant attention, our findings suggest that only a minority of those who accept TasP’s effectiveness express hesitance to rely on it. Actual use of TasP (the final stage of adoption) may be less common because it depends on having partners of different HIV status. Nevertheless, 30.2% of participants who were willing to rely on TasP also reported having engaged, in the prior six months, in condomless sex with a serodifferent partner while the person with HIV had an undetectable viral load, indicating that willingness was not purely speculative. These results suggest that once people understand TasP’s effectiveness, progression through the remaining stages of adoption may be more readily achieved.

Our findings are limited by the use of a convenience sample and may not be generalizable to all U.S. men and TGNCNB individuals who have sex with men. Nonetheless, convenience sampling remains one of the most accessible approaches for studying sexual and gender-minority populations, and results from our relatively large sample are consistent with other surveys on TasP.^4,6–8^ Because participants self-enrolled via advertisements on social media and dating or hookup apps, our findings might be more representative of individuals who use these platforms and are willing to volunteer for public health research. These recruitment strategies may also lead to underrepresentation of people in lower socioeconomic status, from non-urban areas, and of racial/ethnic identifies other than White and not Latino. The cross-sectional design precludes causal inference. In the absence of validated measures of TasP acceptability, we developed survey items based on prior research, which may limit measurement precision. Self-reported data are subject to recall error and respondent fatigue and, while social desirability bias is likely minimal in self-administered confidential surveys, some participants may have given responses that they perceived as more acceptable than their true beliefs.

## Conclusions

Despite these limitations, our study provides a comprehensive view of all stages of TasP adoption among a large sample of U.S. men and TGNCNB individuals who have sex with men, whereas most prior research has examined only selected aspects. Our findings suggest that the main threshold in the TasP adoption cascade is the point at which people move beyond awareness of the strategy to understanding and accepting its effectiveness. While TasP adoption may increase over time as information becomes more widespread, promotion efforts can help reach groups who lag in the process, such as HIV-negative individuals who have never used PrEP. Reaching out to individuals who are not already connected to sexual-health care services could be done via online meeting platforms or other community spaces. LGBTQ communities of color should be prioritized, and educational interventions should not presume familiarity with complex scientific concepts and anticipate skepticism. Healthcare providers also play a central role in deepening TasP knowledge among their patients and should receive appropriate training to facilitate this process. Educating community members and patients should not only involve giving information about TasP, but discussing the strategy and how it can be used effectively. Strengthening confidence in TasP’s effectiveness will help the strategy realize its potential to reduce both HIV transmission and HIV-related stigma.

## Declarations

### Ethics Approval

The study presented in this article was approved by the Institutional Review Board at Columbia University Medical Center (protocol AAAS8251). Data analyses presented in this article were also approved by the Institutional Review Board at the New York City Department of Health and Mental Hygiene (protocol 24-046).

### Consent

All individuals provided informed consent prior to participating in the study presented in this article.

### Data Availability

Data from this study are not publicly available. Please contact the authors with specific inquiries.

### Authors’ Contribution

All authors contributed to the study conception and design, material preparation, data collection, and analysis. The first draft of the manuscript was written by the lead author and all authors commented on previous versions of the manuscript. All authors read and approved the final manuscript.

## Supplemental material

*The HIV-treatment-as-prevention adoption cascade among U.S. men and gender-minority individuals who have sex with men*

## Measures of TasP adoption and TasP-related behaviors

### Awareness of TasP/U=U

To evaluate awareness of TasP/U=U, we presented participants with the following statement and question:

The next series of questions will ask what you know and think about HIV treatment as prevention (TasP), which is also known as *Undetectable = Untransmittable* or *U=U*.

According to the U.S. Centers for Disease Control (CDC), TasP or U=U is defined as the following: A person with HIV who takes HIV medicine as prescribed and gets and stays virally suppressed or undetectable has effectively no risk of sexually transmitting HIV to HIV-negative partners.

Prior to reading this, did you know about TasP or U=U?

Answer choices were:

1. No, I had never heard of TasP/U=U,
2. I had heard about TasP/U=U but didn’t really know what it meant, and
3. Yes, I knew about TasP/U=U.

For this analysis, we categorized participants who said they had previously heard about TasP/U=U (answers 2 and 3) as being aware of TasP/U=U.

### Perceived effectiveness of TasP/U=U

We asked participants to rate the effectiveness of different HIV prevention strategies with the following question:

Phillip is a cisgender man who is HIV positive. If he has anal sex as the top with an HIV-negative partner, what would be the risk of HIV transmission in each of the following situations?

a. Phillip is not undetectable. No condoms are used. The HIV-negative partner is not taking PrEP.
b. Phillip is not undetectable. He wears a condom. The HIV-negative partner is not taking PrEP.
c. Phillip is not undetectable. No condoms are used. The HIV-negative partner is taking PrEP.
d. Phillip is undetectable. No condoms are used. The HIV-negative partner is not taking PrEP.

For each prevention method, participants could select the following choices:

1. Absolutely no risk
2. Very low risk
3. Low risk
4. Moderate risk
5. High risk
6. Very high risk
7. Absolute risk

For this analysis, we looked at participants’ responses on the item related to TasP: “a. Phillip is undetectable. No condoms are used. The HIV-negative partner is not taking PrEP.” Although there is effectively no risk of HIV transmission in this scenario, “statistically a non-zero risk estimate can never be completely ruled out.” Therefore, we considered participants to perceive TasP as effective if they selected 1 (Absolutely no risk) or 2 (Very low risk) to this item.

### Willingness to engage in TasP-related sexual behavior

The survey included series of items to assess participants’ willingness to engage in condomless penetrative sex as the top/insertive partner and as the bottom/receptive partner with individuals of various HIV and ART/PrEP use status. The two series of items were prefaced with the following:

How willing are you to be penetrated by (to bottom for) partners of the following HIV status without a condom, if they have no other sexually transmitted infections? (They insert their penis in your anus, vagina, or front genital opening without using a condom.)

How willing are you to penetrate (to top) partners of the following HIV status without a condom, if they have no other sexually transmitted infections? (You insert your penis in their anus, vagina, or front genital opening without using condoms.)

For each question, participants were asked to rate their willingness to engage in the behavior with the following five types of partners:

a. HIV-negative partner not taking PrEP
b. HIV-negative partner taking PrEP
c. HIV-positive partner who is not undetectable
d. HIV-positive partner who is undetectable
e. Partner whose HIV status I don’t know

For each behavior and type of partner, participants could select the following:

1. Completely unwilling
2. Somewhat unwilling
3. Somewhat willing
4. Completely willing

For this analysis, we considered HIV-negative participants to be willing to engage in TasP-related sex if they selected being completely or somewhat willing to have condomless sex either as top or bottom for an HIV-positive partner who is undetectable. We considered HIV-positive participants to be willing to engage in TasP-related behaviors if they indicated being completely or somewhat willing to have condomless sex either as top or bottom for an HIV-negative partner who is not taking PrEP.

### Recent engagement in TasP-related sexual behaviors

Participants were asked the following four questions about their sex partners in the prior 6 months:

How many sex partners have you had in the past 6 months (since [date 6 months ago automatically generated])? (With how many different people have you have any kind of sex with, e.g., oral, anal, mutual masturbation, fetish play, etc.?)

With how many of your [number entered to question above] partner(s) from the last 6 months did you have penetrative sex? (I.e., anal, vaginal, front hole, whether as top or bottom)

You said you had penetrative sex with [number entered to question above] partner(s) in the past 6 months. With how many of them did you do so without using condoms?

Of what HIV status were the [number entered to question above] partner(s) with whom you had condomless sex in the past 6 months? (Check all that apply.)

a. Partners I knew were HIV negative and taking PrEP
b. Partners I knew were HIV negative but not taking PrEP
c. Partners I knew were HIV negative but I didn’t know their PrEP use
d. Partners I knew were HIV positive and undetectable
e. Partners I knew were HIV positive and whose viral load I knew was detectable
f. Partners I knew were HIV positive but couldn’t be sure of their undetectable status
g. Partners whose HIV status I didn’t know

For this analysis, we considered participants to have engaged in TasP-related sex if they self-reported being never being diagnosed with HIV and checked that they had condomless sex with partners who were HIV positive and undetectable (d). We excluded participants with HIV who did not report being virally suppressed, and considered those with undetectable HIV to have engaged in TasP-related sex if they checked that they had condomless sex with partners who were HIV negative but not taking PrEP (b).

